# Computational modelling of cardiac perfusion to guide Percutaneous Coronary Intervention: a treatment planning tool

**DOI:** 10.1101/2025.10.06.25337398

**Authors:** Giovanni Montino Pelagi, Riccardo Maragna, Giovanni Valbusa, Silvia Bertoluzza, Gianluca Pontone, Christian Vergara

**Affiliations:** Laboratory of Biological Structure (LaBS), Department of Chemistry, Materials and Chemical Engineering (DCMC) at Politecnico di Milano, Milano; Perioperative Cardiology and Cardiovascular Imaging Department at Centro Cardiologico Monzino, Milano; Bracco Imaging S.p.A., Milano; IMATI, Consiglio Nazionale delle Ricerche, Pavia

**Keywords:** computational modeling, coronary artery disease, coronary revascularization, myocardial ischemia, percutaneous coronary intervention, virtual pci

## Abstract

**Objective:** treatment of obstructive coronary artery disease (CAD) requires accurate planning to ensure effective revascularization and full restoration of myocardial perfusion. In this study, we introduce Virtual PCI, a novel computational tool designed to support pre-operative planning of Percutaneous Coronary Intervention (PCI) by predicting the hemodynamic consequences of selected revascularization treatments.

**Methods:** the tool leverages a fully personalized 3D multiscale perfusion model, calibrated using pre-intervention stress CT perfusion (CTP) imaging, to simulate the hemodynamic impact of different revascularization strategies in terms of post-intervention stress myocardial blood flow (MBF) and FFR. The computational framework is also capable of computing the FFR index. We conduct a validation study on patients treated with elective PCI and compare model predictions with dynamic stress CTP at follow-up.

**Results:** the validation study demonstrates high accuracy in predicting post-PCI myocardial perfusion, including potential residual ischemia and cardiac mass at ischemic risk. Through an integrated analysis with FFR, the tool shows potential for its prospective use, identifying in two patients optimal treatment strategies and, in one case, outperforming the executed revascularization in reduction of ischemic burden.

**Conclusions:** Virtual PCI enables the prediction of post-PCI myocardial blood flow (MBF) and FFR, offering a comprehensive assessment of treatment outcomes to identify the best revascularization option from the hemodynamic standpoint.

**Significance:** since it relies solely on non-invasive imaging (cCTA, stress-CTP), Virtual PCI can be integrated early in the diagnostic workflow, providing cardiologists with a powerful, patient-specific tool to optimize PCI planning.

## INTRODUCTION

**E**FFECTIVE treatment of Coronary Artery Disease (CAD) is a delicate clinical matter, which requires an accurate risk stratification of patients integrating information from multiple diagnostic modalities to define the optimal therapeutic option. In patients with obstructive CAD, where the buildup of atherosclerotic plaque narrows the arterial lumen leading to a flow-limiting stenosis, the recommended treatment is usually Percutaneous Coronary Intervention (PCI) [1], consisting on the restoration of coronary lumen patency through balloon inflation.

The FAME clinical trials [2], [3] established important guidelines for the procedure, showing that PCI guided by the Fractional Flow Reserve index (FFR) leads to the best clinical outcomes. However, choosing which and how many lesions to treat is still difficult in many scenarios, including multi-vessel disease, diffuse disease and sequential stenoses [4]. These complex conditions make it exceptionally hard to optimize an intervention that guarantees recovery in FFR value and sufficient restoration in blood perfusion at the tissue level. In this context, *in silico* methods can provide invaluable information, virtually performing PCI procedures to predict their effects on hemodynamics in advance. This idea has been successfully applied to predict FFR response to stenting in an interventional setting, based on invasive angiographic images [5], [6], and in a pre-intervention setting, using coronary CT angiographic scans [7], [8]. However, there are no studies focusing on the prediction of post-revascularization Myocardial Blood Flow (MBF) at the cardiac tissue level, despite its high prognostic value [9]. 3D finite element perfusion models have recently shown incredible potential for personalization through the integration of functional data, even if outside the context of PCI [10]–[14], becoming the most appealing candidates for this approach. Another examples of virtual planning of stenting procedures can be found for the carotid artery [15].

In this work we present, for the first time at the best of our knowledge, a computational tool designed to predict the outcomes of PCI procedures in terms of both hemodynamics in the large coronaries and MBF in the cardiac tissue. Starting from our 3D multiscale perfusion model presented in [16], we introduce a personalized calibration of microcirculation parameters using pre-revascularization MBF maps from dynamic stress-CT perfusion imaging (stress-CTP). We then propose a protocol to virtually simulate PCI, incorporating virtual lumen restoration in the 3D geometry of the coronary arteries and adaptation in model parameters related to the distal resistances.

We validate our framework in a cohort of six patients for a total of 15 treated lesions through direct comparison of the predicted post-operative MBF with stress-CTP at follow-up, and we also explore its use in a prospective fashion.

Our work paves the way to the computational optimization of PCI procedures, for three main reasons: i) it enables the possibility to simulate different treatment options (in terms of which and how many lesions to treat) to identify the one leading to the best hemodynamic results, ii) the hemodynamic analysis is comprehensive, from large coronaries down to tissue-level MBF, and iii) invasive angiographic images are not required, enabling the full pre-operative planning at a very early stage in the clinical path.

## II. METHODS

Figure 1 illustrates the Virtual PCI method, as we envision it for the optimization of coronary revascularization procedures.

**Fig. 1.**
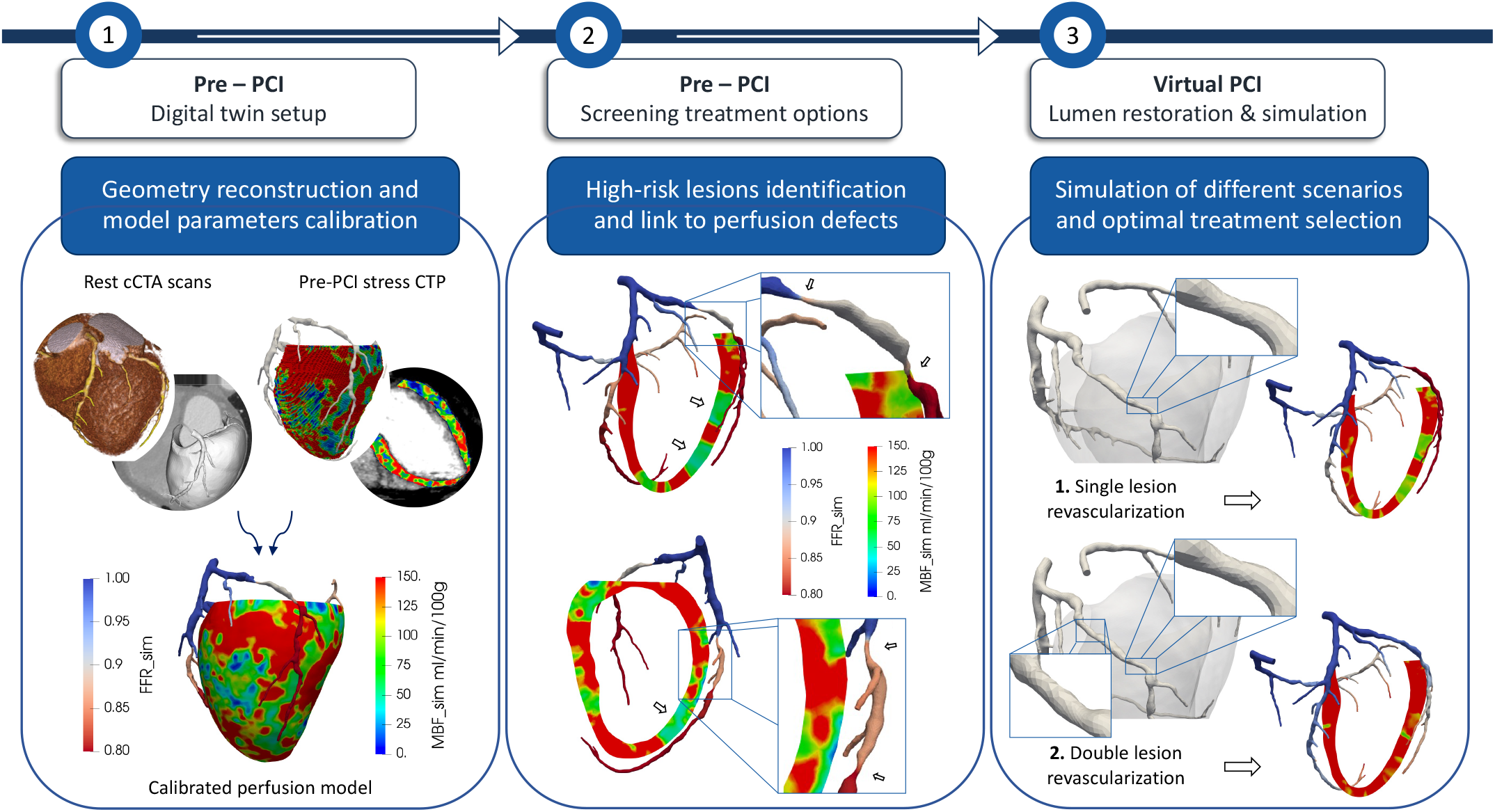
Virtual PCI protocol for clinical usage. 1. Setup of the pre-revascularization calibrated model: reconstruction of the 3D geometries (myocardium and coronary arteries) and calibration of the model parameters through integration of stress-CTP data. 2. Use of the calibrated model for the identification of flow-limiting stenoses, with associated perfusion defects, and screening for potential revascularization strategies. 3. Simulation of different revascularization strategies and selection of the optimal treatment option.

In brief, the Virtual PCI method consists of three steps:

1) setup of the pre-intervention calibrated model through anatomical reconstruction from coronary Computed Tomographic Angiography (cCTA) scans at rest plus the calibration of model parameters using stress Computed Tomographic Perfusion (stress CTP). The calibrated model obtaineid this way represents the physiological and functional state of the patient prior to any revascularization;

2) use of the calibrated model to identify hemodynamically significant coronary lesions (FFR *<* 0.8) which can be associated to perfusion defects in the myocardium (MBF *<* 101 ml/min/100g [18]). Through this step, a screening of the possible revascularization strategies is performed and a first treatment plan is formulated as follows: for each main coronary branch, revascularization is indicated for the most hemodynamically significant lesion that is located upstream to all the perfusion defects identified for that branch;

3) virtual replica of the revascularization procedure by modifying the coronary anatomy to restore lumen patency at the selected treatment sites, plus post-intervention hemodynamics simulation and assessment of post-intervention FFR and MBF maps. Should any of the perfusion defects persist, for example due to the presence of untreated lesions in the same coronary branch, the computational process is repeated with the identification of new treatment sites, until the highest restoration of MBF is achieved across the entire myocardium.

The optimized revascularization strategy suggested by our method, therefore, consists of all the treatment sites identified this way and is provided to the clinicians before the interventional stage is reached, so that an optimal procedure can be established based on hemodynamic and technical considerations.

In the following sections we detail the methodology of each step: in Section II-A we provide the details on image acquisition protocols and anatomical reconstruction, in Section II-B we provide information on the computational framework used, whereas in Section II-C we discuss the core of the Virtual PCI approach, including the pre-intervention calibration strategy, how we virtually replicate the revascularization intervention and how we simulate the post-intervention hemodynamics.

Finally, in Section II-D we provide the methods of a validation study performed on 6 patients from Centro Cardiologico Monzino, including population characteristics and inclusion/exclusion criteria, workflow and validation metrics.

### A. Image acquisition and anatomy reconstruction

On each patient, the two imaging modalities (i.e. rest cCTA and stress CTP) are acquired sequentially in a unified protocol. Before the rest scan, all patients received sublingual nitrates to ensure coronary vasodilation. Rest cCTA scans were obtained as first-pass scans, while stress-CTP scans were acquired after the rest protocol and after vasodilation induced by an intravenous adenosine injection (0.14 mg/kg/min over 4 min). The dynamic protocol consisted in ECG-gated acquisition over multiple (*≃* 25-30) heartbeats to enable the quantitative analysis of the contrast medium kinetics, allowing MBF computation. All details on scanner and detector technology, acquisition parameters, contrast medium administration and image reconstruction can be found in [17] and [18]. Follow-up imaging were obtained using the same methodology, 5-6 years after revascularization.

3D segmentations of the patient’s myocardium and coronary arteries are performed by us on pre-intervention resting cCTA images. The myocardium segmentation is obtained automatically using the TotalSegmentator tool of 3D Slicer [19], whereas the lumen of the coronary arteries is semiautomatically segmented using the level set segmentation algorithm in VMTK [20]. Since the specific HU values of each image depend on the particular processing algorithm used by the CT scanner at the acquisition stage, the threshold values for segmentations are selected as follows: i) high threshold is set to 100 HU higher than the HU sampled at the aortic root. This choice excludes the high-density atherosclerotic plaques including calcifications; ii) low threshold is picked branch by branch following visual inspection, so that only the lumen is segmented excluding the surrounding tissue as well as low-density atherosclerotic plaques including fibrous and lipidic components. 3D segmentations of the coronary arteries are pre-processed using a smoothing filter to achieve a final surface of sufficient quality to be used in computational simulations. The left myocardium is meshed in VMTK using tetrahedral elements with target edgelength of 1.5 mm. The computational mesh for the coronary artery is obtained in VMTK using tetrahedral elements with an adaptive edgelength based on the local distance to the centerline, i.e. an approximated local radius.

### B. Computational framework

Simulations of the hemodynamics in the entire coronary tree, from the epicardial arteries to the capillaries embedded in the myocardial muscle, are performed using our multiscale computational model, presented in [16]. This model features a 3D incompressible fluid-dynamics description of blood flow in the epicardial arteries coupled with a 3D multicompartment Darcy model for the microcirculation.

The Darcy formulation we use is a special version, which accounts for the compliance of microvessels and a time-varying intramyocardial pressure surrogating heart contraction, so that the whole heartbeat can be successfully simulated.

The two models are coupled through interface conditions that enforce mass conservation and balance of interface forces, as firstly used in [21]. Geometrical coupling is achieved through the subdivision of the myocardium into perfusion regions and a one-to-one association between each coronary branch and a single perfusion region, as previously described [21].

We report here the formulation of the coupled multiphysics model:

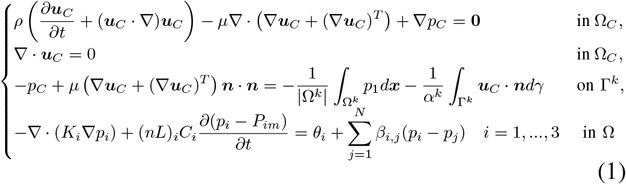

The first two lines in 1 are the incompressible Navier-Stokes formulation: *ρ, µ*, ***u***_*C*_ and *p*_*C*_ are the blood density, viscosity, velocity and pressure in the epicardial arteries, respectively.

This formulation is complemented with the standard no-slip condition at the coronary wall and with a patient specific pressure waveform as inlet boundary condition. This patient specific pressure inlet BC is built following the methodology described in [22] starting from basic measures of heart rate and brachial systolic and diastolic pressures.

The fourth line in 1 is a set of three Darcy equations (multi-compartment Darcy) with the underscript *i* denoting a specific compartment: *p* is the unknown pore pressure, *K*, (*nL*), *C* are the comprtment specific permeability, length density and compliance respectively, and *β*_*i,j*_ is the inter-compartment conductance between compartments *i* and *j*. The source terms *θ*_*i*_ represent the mass source and sink terms given by:

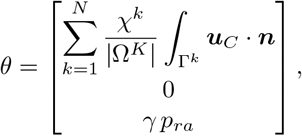

with:

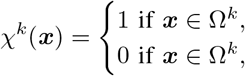

The first line is the source term representing blood coming from the epicardial coronaries and the third line is the sink term accounting for venous return, with *γ* being the venous drain coefficient *p*_*ra*_ the right atrium pressure.

The third line in 1 is the interface condition coupling the Navier-Stokes and Darcy problems, prescribing, at each coronary outlet Γ^*k*^, a flow proportional to difference between blood pressure at said outlet and the average pressure of the first Darcy compartment in the corresponding perfusion region Ω^*k*^. The relationship between this pressure difference and the outlet flow is regulated by the parameter *α*^*k*^, which represents the conductance of the coronary portion between the 3D outlet and the first Darcy compartment. This coupling condition for the normal component of the force is complemented with null tangential tractions for the two tangential directions.

The multiphysics model 1 is solved through a strongly coupled, partitioned scheme where the NS and Darcy sub-problems are solved independently and iteratively at each time step, until convergence. Details on the numerical implementations can be found in [22].

All the numerical parameters and the majority of physics-related parameters have been optimized in a previous study [16] using population-based blood flow measures and experimental measures. In this work we use the same parameter values as reported in [16], with the only exceptions of the distal conductances *α*^*k*^ and the microcirculation length densities of vessels (*nL*)_*i*_: for these parameters we take a further step forward and we propose a personalized calibration based on perfusion imaging, as discussed in detail in Section II-C.

All numerical simulations are run using the software life^x^, a high performance library for Finite Elements simulations of multiphysics, multiscale and multidomain problems developed at MOX - Dipartimento di Matematica, in cooperation with LaBS - Dipartimento di Chimica, Materiali e Ingegneria Chimica, both at Politecnico di Milano [23], [24].

### C. Virtual PCI approach

To achieve the *in silico* replica of the patient at the pre-intervention stage and to simulate the post-intervention hemodynamics we use the following approach:

1) initial simulation with semi-personalized model calibration. After anatomical reconstruction and meshing, we perform a *base* simulation using the following semi-personalized model calibration:

- inflow boundary conditions are prescribed as customized pressure waveforms at the aortic root, built based on the patient’s basic information (age, sex, height, weight) and routine clinical measures (heart rate, brachial systolic/diastolic pressure) as explained in [22];
- microvascular parameters in the Darcy model are taken as constants over space, with the values reported in our previous study [16];
- distal conductances of the epicardial coronary branches are calibrated using an angiographic criterion based on the radiological evaluation of the cCTA scan, following a concept introduced in a previous study [25]. For each branch outlet *k*, the corresponding distal conductance *α*^*k*^ is modulated based on the presence and severity of the stenoses in that branch. We set a physiological value of *α*^*k*^ = 1 *\**10^*−*10^ m^3^ · s^*−*1^ · Pa^*−*1^ for all branch outlets where no or only minimal lesions (*<*= 30% stenosis) are documented in the upstream path from said outlet to the aortic root. In the case of moderate, severe and critical lesions we set the value of *α*^*k*^ according to Table I, where all the scores result from clinical evaluation of cCTA scans performed by expert cardiologists. For example, in the case of a severe stenosis in the medial portion of the Left Anterior Descending (LAD) artery, all the outlets located distally to said lesion have their distal conductance set to *α*^*k*^ = 0.25 *\**10^*−*10^ m^3^ ·s^*−*1^ ·Pa^*−*1^, whereas the outlets located proximally with respect to the lesion have the physiological value of *α*^*k*^ = 1*\**10^*−*10^ m^3^ · s^*−*1^ · Pa^*−*1^. In the event of multiple lesions affecting the same branch, the value of *α* is set according to the lesion with the highest severity;

**TABLE 1.** VALUES ATTRIBUTED TO THE DISTAL CONDUCTANCE ***α***^***k***^ OF EACH BRANCH OUTLET ***k*** ACCORDING TO THE PRESENCE AND SEVERITY OF CORONARY LESION AT ANY POINT IN THE UPSTREAM PATH FROM THE OUTLET TO THE AORTIC ROOT.

| Stenosis degree | Lesion classification | Alpha $\text{m}^3 \cdot \text{s}^{-1} \cdot \text{Pa}^{-1}$ |
| --- | --- | --- |
| < 30% | Minimal | $1 * 10^{-10}$ |
| 30 - 50 % | Mild | $0.75 * 10^{-10}$ |
| 50 - 70 % | Moderate | $0.5 * 10^{-10}$ |
| 70 - 90 % | Severe | $0.25 * 10^{-10}$ |
| > 90% | Critical | $0.1 * 10^{-10}$ |

2) integration of CTP imaging data and fully personalized model calibration. We project the pre-revascularization MBF map obtained by dynamic stress-CTP onto the computational mesh and compute a correction factor for the parameters related to the microvasculature:

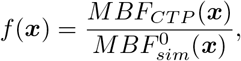

where *MBF*_*CTP*_ is the MBF from dynamic stress-CTP imaging, 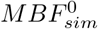 is the MBF computed by the model at step 1 and the notation (***x***) indicates that these quantities are scalar fields that vary in space over the myocardium. We then modulate for each Darcy compartment *i* the local density of vessels (*nL*)_*i*_ using the following formula:

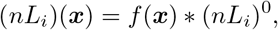

where (*nL*_*i*_)^0^ represent the values of length density (constant over space) used in the *base* simulation;

3) restoration of coronary lumen patency and post PCI simulation. To replicate the post revascularization angiographic state of the treated coronary arteries, the 3D geometry at the stenotic treated sites is inflated, restoring the lumen caliber using as a reference a proximal or distal portion of the branch, considered angiographically normal. This step can be tailored in terms of the entity of lumen restoration and the extent of the vessel interested by the procedure. For example, one can replicate a good angiographic result by fully restoring lumen patency, or replicate residual stenoses, due to elastic recoil phenomena or to some issues during the procedure. Also, the use of stents of different lengths can be replicated by adjusting the length of the stenotic segment interested by the inflation process. After the update of the coronary geometry, the final surface is meshed and used for the numerical simulation of the post-PCI hemodynamic state. In this step, we use the following setup:

- multiphysics cardiac perfusion model reported in II-B with personalized aortic pressure inlet conditions,
- microvascular parameters coming from the fully personalized calibration, in particular the 3D map of local length densities (*nL*)_*i*_(***x***). With this choice we assume that, in the short term after the intervention, microcirculation parameters are not affected by the PCI procedure,
- distal conductances *α*^*k*^ updated according to the post-PCI angiographic state, following the criteria in Table I. In the case of residual stenoses after the treatment, those are still incorporated into the model using this approach.

### D. Validation study

The capability of the Virtual PCI method to quantitatively predict the hemodynamic effects of a specific revascularization procedure is established through a retrospective human validation study.

Inclusion criteria for the study were: enrollment in the PERFECTION clinical study [17] (with associated inclusion and exclusion criteria, briefly: patients with intermediate to high risk of CAD without previous history of myocardial infarction, revascularization procedures and/or emergency procedures), execution of rest cCTA and dynamic stress CTP imaging, execuction of elective PCI treatment and of follow-up dynamic stress-CTP imaging. Exclusion criteria were: non-diagnostic quality of cCTA or CTP imaging, significant (*>* 50%) in-stent restenosis in patients treated with stents, significant disease progression in any major coronary between the time of PCI and the follow-up CTP imaging. The final population count *n* = 6 patients and a total of *m* = 15 coronary lesions treated. Figure 2 illustrates the application of the Virtual PCI method in the validation study. After the creation of a calibrated model as previously explained, the specific PCI clinically executed on the patient is virtually replicated by restoring the coronary lumen at the treated sites. In this step, we use invasive angiographic images to ensure that the lumen restoration we apply to the virtual replica matches the one effectively achieved on the patient in the interventional setting, in terms of location of the treated site, level of restoration in lumen patency and length of the treated segment. Lastly, we perform a post-PCI perfusion simulation and we compare the Virtual PCI predictions with dynamic stress CTP at follow-up. Validation is carried out at multiple levels: visual inspection of perfusion defects, evaluation of the amount and distribution of the cardiac mass at ischemic risk and clinical evaluation of the myocardial regions based on the 17 segments AHA model [26]. All the comparisons are performed between the predictions of the Virtual PCI method vs post-PCI stress-CTP imaging collected at follow-up.

**Fig. 2.**
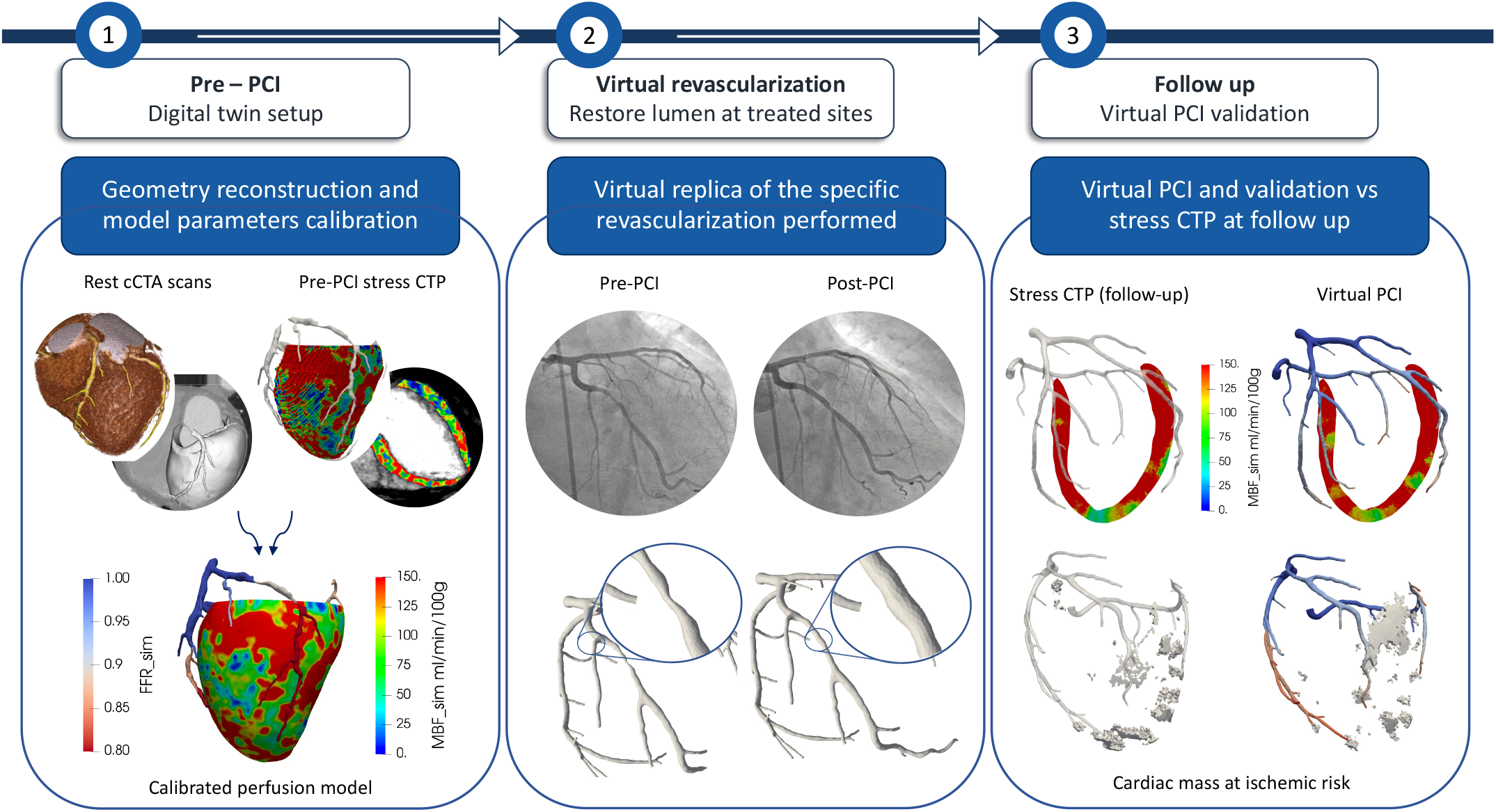
Validation protocol for the Virtual PCI method. 1. Setup of the pre-revascularization calibrated model: reconstruction of the 3D geometries (myocardium and coronary arteries) and calibration of the model parameters through integration of stress-CTP data. 2. Angiography-guided lumen restoration at the treated sites to virtually replicate the specific revascularization performed. 3. Validation with direct comparison between Virtual PCI predictions and follow-up stress CTP imaging.

For the visual inspection validation, representative long-axis slices are oriented after the identification, in the pre-PCI stress CTP, of the regions with the most clinically relevant impairment in blood flow, defined as the largest region with reduced MBF which extends through the entire thickness of the myocardial wall. This is done to match the analysis method currently used in the clinical evaluation of stress CTP, since regions with reduced MBF are considered true perfusion defects if they involve the subendocardial layer of the myocardium (subendocardial defect) or the entire myocardial wall (transmural defect) rather than the subepicardial layer [17]. Visual inspection is then performed using a narrow window for MBF of [0-150 ml/min/100g] [17], [18].

Validation based on the mass at ischemic risk is carried out by extracting, through thresholding, the cardiac mass showing MBF *<* 101 ml/min/100g. This cutoff has been validated in the RIPCORD clinical study [18] based on the quantification of absolute MBF in stress conditions, which represents an independent indicator of the cardiac mass with inducible ischemia [27]. We recall that, for a given region in the myocardium, the condition of inducible ischemia implies the absence of actual ischemia while the patient is in resting conditions and, however, the insurgence of acute ischemia in the event of an increased metabolic demand, such as during physical activity. The cardiac mass at ischemic risk in the follow-up stress CTP images is extracted from the mass that showed inducible ischemia already at the pre-intervention stage. Specifically, we apply a first thresholding for pre-PCI MBF *<* 120/min/100g and then a second thesholding on follow-up MBF *<* 101 ml/min/100g. In this step, we propose the use of a slightly larger cutoff value of 120 ml/min/100 for the pre-PCI thresholding to also capture the cardiac mass very close the regions classified as “at ischemic risk”. This prevents ass at risk underestimation in the follow-up imaging.

Clinical validation is performed in each patient based on the classification, performed by an expert cardiologist, of each of the 17 segments of the AHA model [26] according to the presence of subendocardial or transmural perfusion deficits in said myocardial segment. To better differentiate between a high certainty perfusion defect from a defect whose evaluation is more uncertain, we propose the use of the following cutoffs, in accordance with two expert cardiologists:

- Perfusion deficit: presence of a sub-endocardial or transmural region with values of *MBF <* 90 ml/min/100g;
- Borderline impaired flow: presence of a sub-endocardial or transmural region with values of 90 *< MBF <* 105 ml/min/100g;
- Normal perfusion: absence of any sub-endocardial or transmural region with values of *MBF <* 105 ml/min/100g;

*FFR*_*sim*_ values refer to the FFR index computed from the numerical simulation, either at the pre-interventional stage and as output of the Virtual PCI method. The specific values reported in Table II are computed according to:

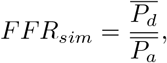

where 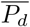 and 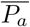 are the time-averaged distal and aortic pressures, respectively. As distal site, we take the a point in the coronary tree distal to all the coronary stesoses, where the coronary artery has diameter *≃* 0.8 mm. In the case of absence of lesions, a default value of *FFR*_*sim*_ = 0.95 is used.

**TABLE 2.**
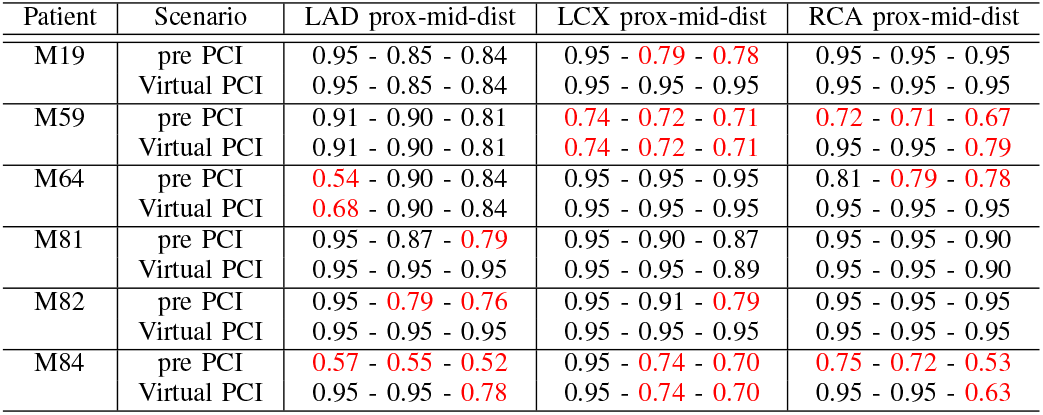
RESULTS OF THE VALIDATION STUDY: ***F F R***_***sim***_ **INDEX**. FOR EACH PATIENT IS REPORTED THE ***F F R***_***sim***_ INDEX BEFORE AND AFTER THE PCI REVASCULARIZATION, AT THE PROXIMAL, MEDIAL AND DISTAL POINTS OF EACH MAJOR CORONARY; THIS MAY INCLUDE THE SECONDARY BRANCHES ARISING FROM EACH PORTION. IN RED ARE HIGHLIGHTED THE PORTIONS WITH FFR ***<* 0.8**, WHERE REVASCULARIZATION IS RECOMMENDED. IN BRANCHES WITHOUT ANY STENOTIC SITE, A DEFAULT VALUE OF FFR = 0.95 IS SET.

| Patient | Scenario | LAD prox-mid-dist | LCX prox-mid-dist | RCA prox-mid-dist |
| --- | --- | --- | --- | --- |
| M19 | pre PCI | 0.95 - 0.85 - 0.84 | 0.95 - 0.79 - 0.78 | 0.95 - 0.95 - 0.95 |
|  | Virtual PCI | 0.95 - 0.85 - 0.84 | 0.95 - 0.95 - 0.95 | 0.95 - 0.95 - 0.95 |
| M59 | pre PCI | 0.91 - 0.90 - 0.81 | 0.74 - 0.72 - 0.71 | 0.72 - 0.71 - 0.67 |
|  | Virtual PCI | 0.91 - 0.90 - 0.81 | 0.74 - 0.72 - 0.71 | 0.95 - 0.95 - 0.79 |
| M64 | pre PCI | 0.54 - 0.90 - 0.84 | 0.95 - 0.95 - 0.95 | 0.81 - 0.79 - 0.78 |
|  | Virtual PCI | 0.68 - 0.90 - 0.84 | 0.95 - 0.95 - 0.95 | 0.95 - 0.95 - 0.95 |
| M81 | pre PCI | 0.95 - 0.87 - 0.79 | 0.95 - 0.90 - 0.87 | 0.95 - 0.95 - 0.90 |
|  | Virtual PCI | 0.95 - 0.95 - 0.95 | 0.95 - 0.95 - 0.89 | 0.95 - 0.95 - 0.90 |
| M82 | pre PCI | 0.95 - 0.79 - 0.76 | 0.95 - 0.91 - 0.79 | 0.95 - 0.95 - 0.95 |
|  | Virtual PCI | 0.95 - 0.95 - 0.95 | 0.95 - 0.95 - 0.95 | 0.95 - 0.95 - 0.95 |
| M84 | pre PCI | 0.57 - 0.55 - 0.52 | 0.95 - 0.74 - 0.70 | 0.75 - 0.72 - 0.53 |
|  | Virtual PCI | 0.95 - 0.95 - 0.78 | 0.95 - 0.74 - 0.70 | 0.95 - 0.95 - 0.63 |

## III. RESULTS

In Section III-A we report the results of the retrospective validation study, based on the metrics exposed in Section II-D, while in Section III-B we discuss a prospective application of the Virtual PCI method on two selected patients.

### A. Results of the validation study

Figures 3 and 4 show the visual inspection of perfusion defects, respectively in four patients treated with single or double PCI revascularization (M19, M59, M64, M82) and in two patients treated with a more complex PCI procedure consisting of four or more treated lesions (M81, M84).

**Fig. 3.**
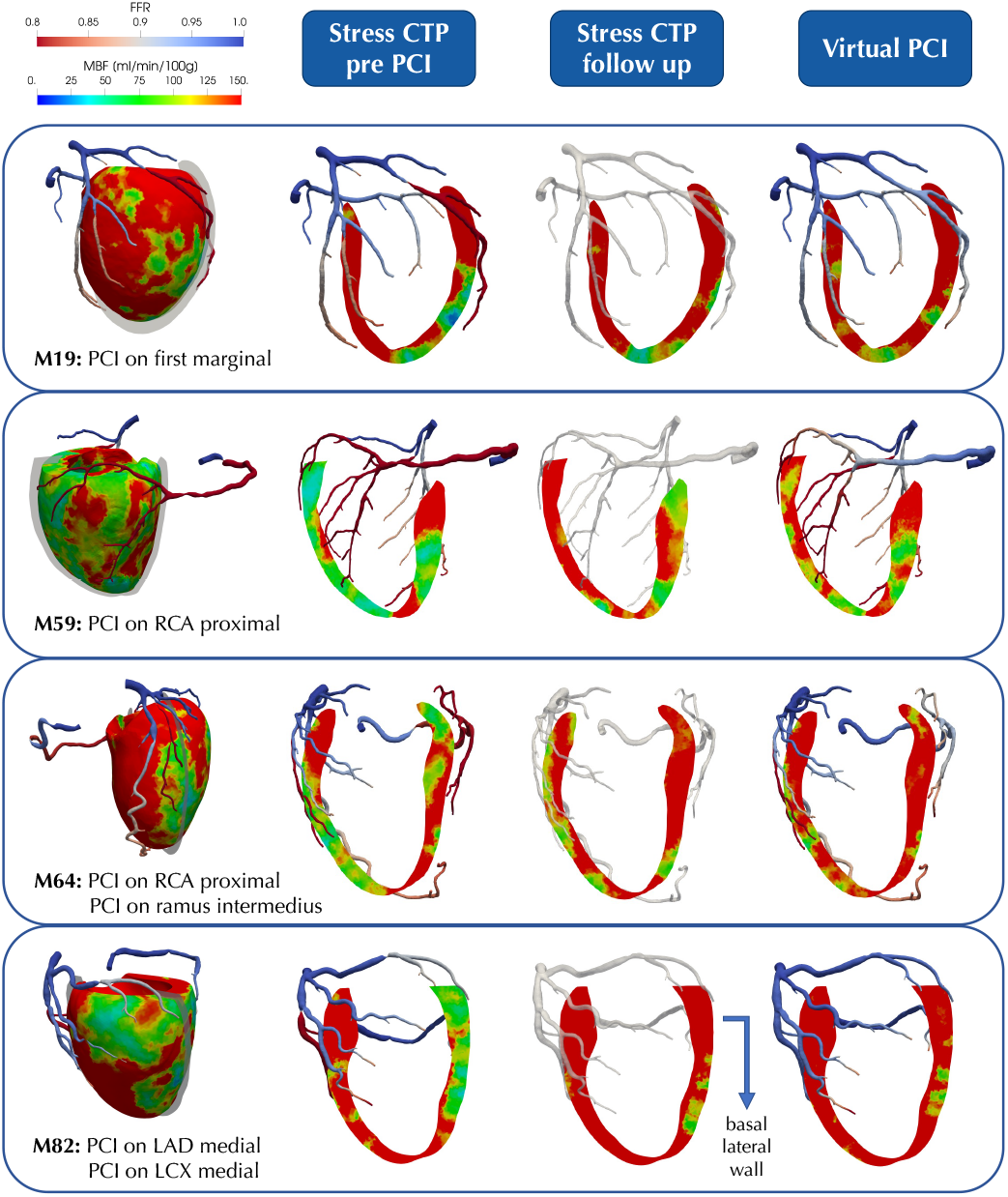
Results of the validation study: patients with single or double revascularization. Each block represents a different patient. Leftmost column: pre-PCI physiological state of the patient obtained with the calibrated perfusion model: FFR index (in the coronaries) and MBF map (in the myocardium); the performed PCI treatment is also specified. Right block of three columns: comparison between dynamic stress CTP maps pre-revascularization (plus FFR index from the calibrated model - left), dynamic stress CTP at follow-up (middle) and Virtual PCI results (right). Virtual PCI results include predicted FFR index and MBF map after revascularization. Visualization slices are aligned with the most significant perfusion defects, as clinically evaluated from pre-revascularization stress CTP.

**Fig. 4.**
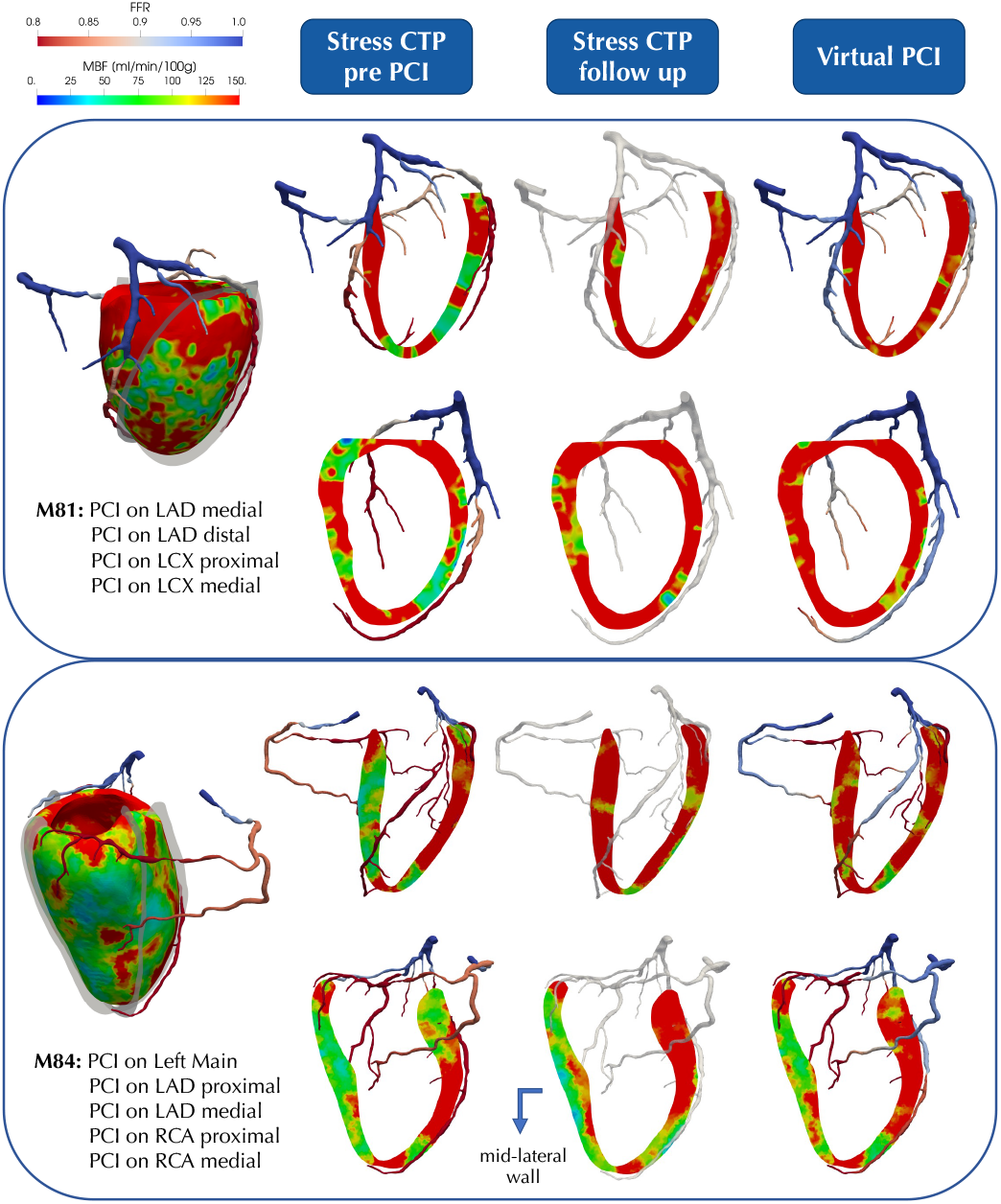
Results of the validation study: patients with complex multi-vessel revascularization. Upper and lower blocks represent two different patients. Leftmost column: pre-PCI physiological state of the patient obtained with the calibrated perfusion model: FFR index (in the coronaries) and MBF map (in the myocardium); the performed PCI treatment is also specified. Right block of three columns: comparison between dynamic stress CTP maps pre-revascularization (plus FFR index from the calibrated model - left), dynamic stress CTP at follow-up (middle) and Virtual PCI results (right). Virtual PCI results include predicted FFR index and MBF map after revascularization. Visualization slices are aligned with the most significant perfusion defects, as clinically evaluated from pre-revascularization stress CTP.

From these results we see that the Virtual PCI method is able to reproduce with good accuracy the evolution of perfusion defects after the restoration of coronary lumen through PCI. Indeed, from the comparison of the second and third columns we see a good agreement, in terms of patterns and values of MBF, between the outcomes of the Virtual PCI simulation and the follow-up CTP, in both groups of patients.

The evolution of perfusion defects is well captured both in the case of complete blood flow restoration, e.g. in the case of the basal lateral wall in patient M82, as well as in the case of incomplete flow restoration, e.g. in the case of the mid-lateral wall in patient M84. These results highlight that, in some cases, there is the possibility for regional perfusion defects to persist even after the treatment of the coronary artery that is associated with that region.

Figure 5 shows the myocardial mass at ischemic risk (MaR), extracted through thresholding as the cardiac mass with *MBF <* 101 ml/min/100g. For each patient, we compute the percentage of mass at ischemic risk and the relative error as:

**Fig. 5.**
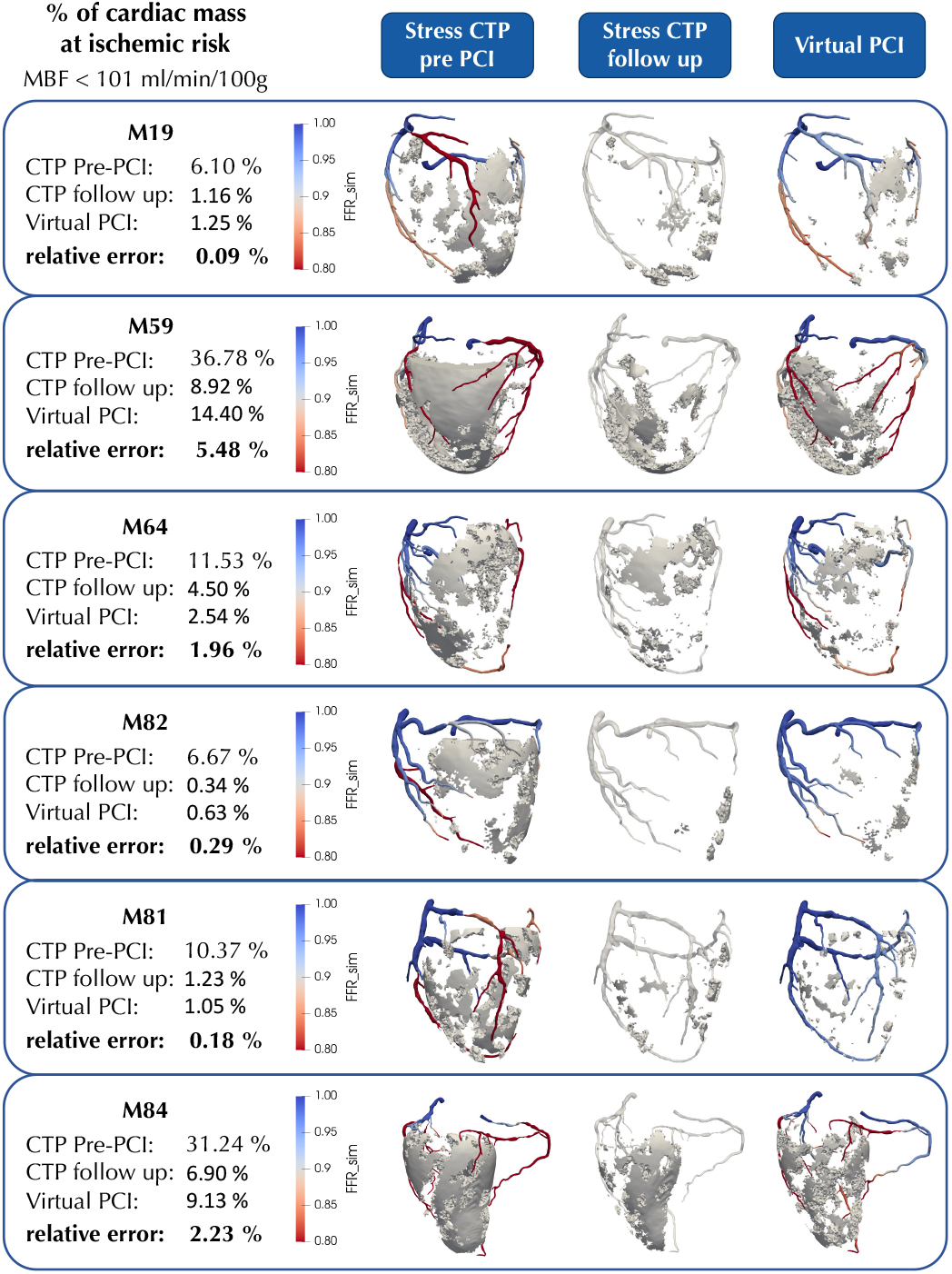
Results of the validation study: cardiac mass at ischemic risk. Each block represents a different patient. Cardiac mass at ischemic risk is extracted through thresholding with a cutoff value of ***MBF <* 101** ml/min/100g. Left column: percentages of mass at risk extracted from pre-PCI stress CTP, follow-up stress CTP and prediction of Virtual PCI method. Right block of three columns: camparison between the spatial distribution of mass at risk extracted from the same sources.

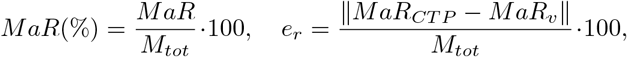

where *M*_*tot*_ is the total myocardial mass, *MaR*_*CTP*_ is the Mass at Risk extracted from the follow-up CTP and *MaR*_*v*_ is the Mass at Risk computed by the Virtual PCI method.

From the results in Figure 5 we see that the Virtual PCI method is successful in quantifying the mass at ischemic risk, with a relative error always below 2.5% for all patients except for M59, where the error is 5.48%, and a mean relative error of 1.7% across all the six patients. From these results, we see that our method shows very high accuracy in the case of a low/moderate amount of cardiac mass at risk prior to PCI. In these cases the restoration of blood flow is captured precisely, whereas we notice a slight overestimation of the mass at risk when the pre-revascularization condition shows extensive inducible ischemia, as in the case of patients M59 and M84.

Figures 6 and Table II summarize the results of the entire cohort of patients from the clinical perspective. In Figure 6 we report the interpretation of perfusion defects based on a personalized version of the 17 segments heart model, which is a well established analysis in clinical practice [26]. For each patient, each segment is attributed to a major coronary artery based on the specific anatomy of the coronary tree. Since the focus of the study is on the myocardial mass perfused by revascularized arteries, we consider out of scope all the segments that are attributed to coronary branches not interested by the PCI procedure. This may include some basal segments when the revascularization is performed at a medial or distal site of the associated artery, for example as in the LAD territory of patient M81. For all the other segments, we assign a color-coded score according to the thesholds defined in Section II-D.

**Fig. 6.**
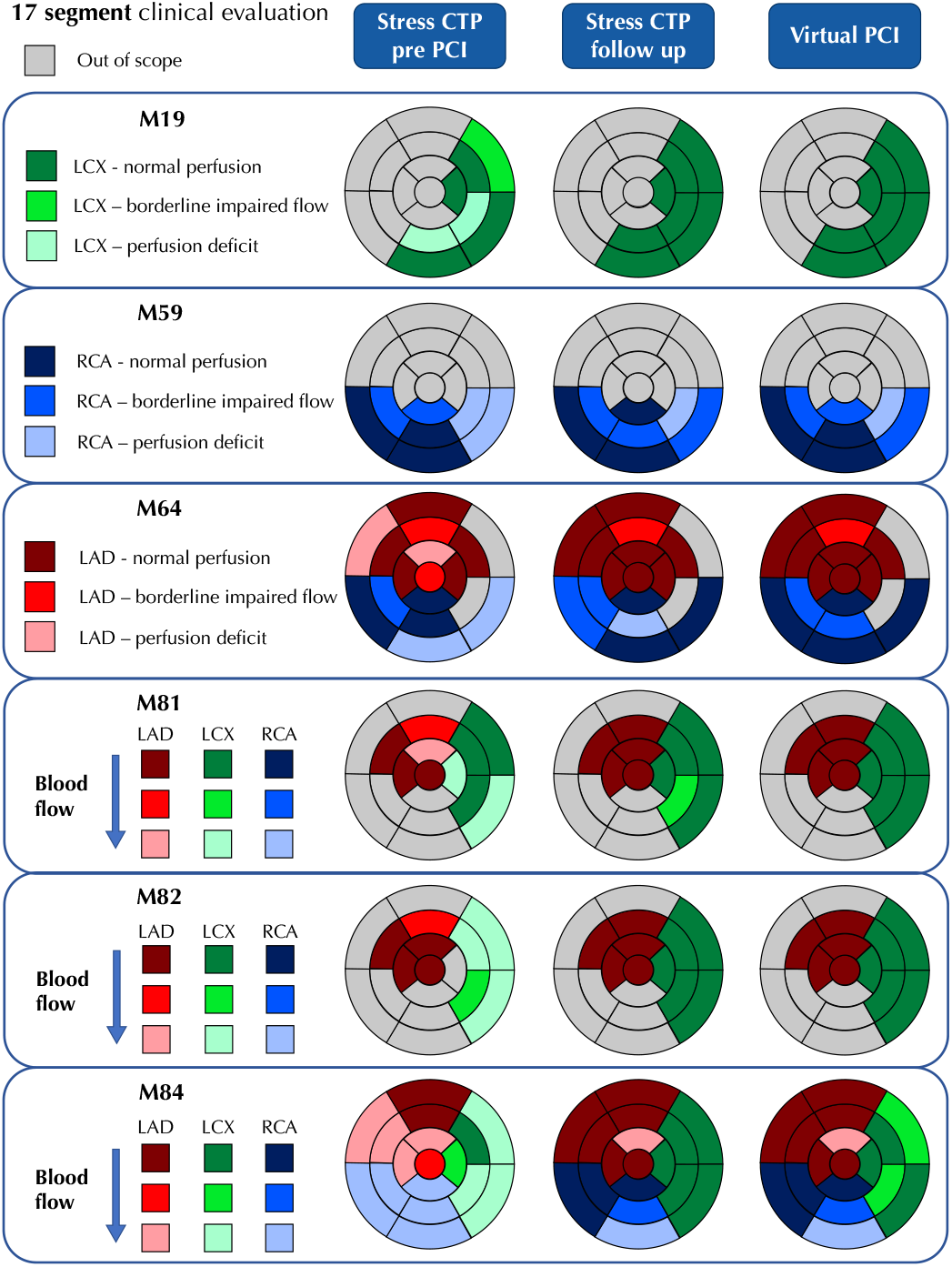
Results of the validation study: 17 segments analysis. Each block represents a different patient. Perfusion deficit: presence of a subendocardial or transmural region with ***MBF <* 90** ml/min/100g; borderline impaired flow: presence of a subendocardial or transmural region with **90 *< MBF <* 105** ml/min/100g; normal perfusion: absence of any subendocardial or transmural region with ***MBF <* 105** ml/min/100g. Segments are marked as out of scope if they are associated with untreated coronary arteries.

The comparison between follow-up CTP imaging and the predictions of the Virtual PCI method in Figure 6 highlights the overall effectiveness of our method in reproducing complete and incomplete restoration of blood flow following PCI treatment. Out of a total of 11 macro-territories affected by PCI (a macro-territory is defined as the union of all the segments related to one of the three main coronary artery - LAD, LCX and RCA), we report perfect segment-wise correspondence between follow-up CTP and Virtual PCI in 7 cases. In two of the remaining cases, both involving the right coronary artery in patients M59 and M64, the Virtual PCI method correctly detects regions of reduced blood flow, showing, however, lower accuracy in the precise localization of the perfusion deficit. Regarding the last two cases, we report slight discrepancies in the LCX territories of patients M81 and M84, with incorrect classification of one and two segments, respectively. Importantly, there are no segments where follow-up imaging documents a perfusion deficit that is completely missed by the Virtual PCI method, or where a normally perfused segment (according to follow-up CTP) is indicated as perfusion deficit by our method.

The results of the 17 segments analysis shown in Figure 6 are particularly interesting when combined with the large artery hemodynamics results shown in Table II, where we report the *FFR*_*sim*_ index computed by the pre-PCI calibrated model and by the Virtual PCI method, the latter representing the physiological state after the restoration of coronary lumen patency.

Out of the six macro-territories where at least one segment is marked as “borderline impaired flow” or “perfusion deficit” after PCI, four are associated with a flow-limiting stenosis, with *FFR <* 0.8, which persists even after the treatment. This occurs in the case of the RCA macro-territory of patients M59 and M84, and the LAD macro-territory of patients M64 and M84. These results indicate that revascularization at specific site can lead to the insurgence of other flow-limiting stenoses in the same coronary branch.

### B. Prospective use of the Virtual PCI method

The prospective utility of the Virtual PCI method in clinical practice is explored in a comparative study. In two selected patients, we apply the Virtual PCI method to simulate different revascularization options with respect to the one clinically executed and we compare the results with the outcomes of the clinally executed PCI. We analyze here the following two scenarios:

**1) Patient M81, minimal PCI:** revascularization at one site for each diseased artery, with treatment sites selected as the most hemodynamically significant lesions located upstream to all the perfusion defects in the myocardium. This scenario represents a reduced version with respect to the PCI clinically executed, with fewer lesions treated;

**2) Patient M84, optimized PCI:** revascularization at all severe and critical stenotic sites, aimed at the restoration of MBF in the entire myocardium. This scenario represents an augmented version with respect to the PCI clinically executed, with more lesions treated.

Figure 7 shows the results of the comparative study, including visual inspection evaluation, mass at ischemic risk quantification and segmental perfusion evaluation. These results highlight the value of using the Virtual PCI method as a pre-operative planning tool for PCI, since the hemodynamic predictions on the same patient vastly differ depending on the the specific PCI procedure, in terms of number and location of stenotic sites treated with revascularization.

**Fig. 7.**
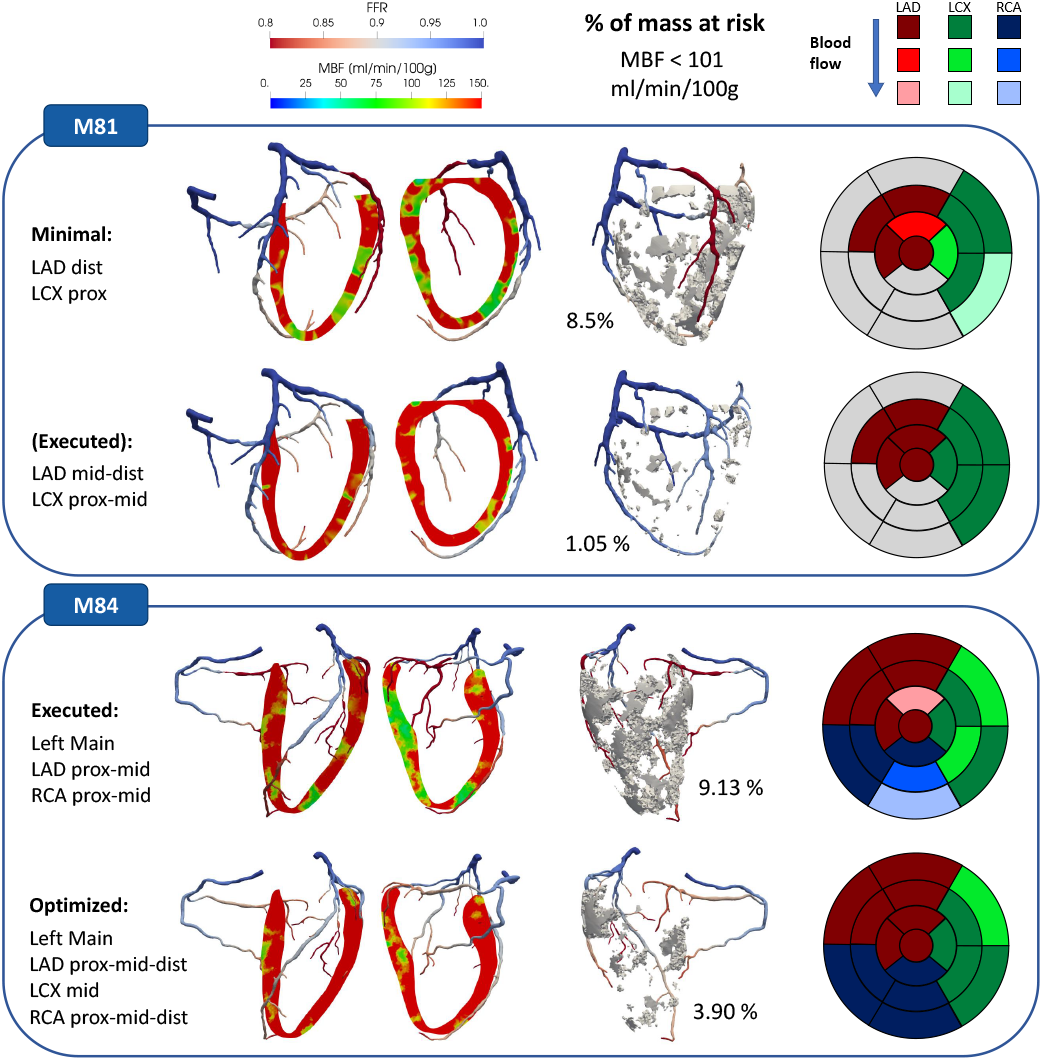
Example of prospective use of the Virtual PCI method. Upper and lower blocks represent different patients; for both are reported the results of the Virtual PCI method applied to two possible revascularization procedures, with the full analysis (including visual inspection evaluation, mass at ischemic risk quantification and segmental detection of ischemia).

In patient M81, where two major coronaries (LAD and LCX) are affected by atherosclerotic disease resulting in two sequential stenoses on each branch, a hypothetical PCI scenario of minimal revascularization (treating only one lesion per artery) shows an incomplete restoration of MBF in the myocardium, resulting in a high amount of mass at ischemic risk (*≃*8.50%) even after the procedure and in the persistence of perfusion defects in multiple segments. This is particularly evident in the LCX territories and is further corroborated by the post-PCI *FFR*_*sim*_ results, showing that, in the absence of the distal lesion in the LCX, the remaining lesion in the mid segment of the artery has now flow-limiting behaviour (*FFR*_*sim*_ *<* 0.8). In the context of a prospective use of the Virtual PCI method, therefore, this specific PCI procedure would be contraindicated based on its predicted hemodynamic outcomes.

In patient M84, where the diffuse atherosclerotic disease leads to multiple stenotic sites, the post-PCI hemodynamic state computed by the Virtual PCI method following the clinically executed procedure shows residual perfusion defects across multiple segments (confirmed by the follow-up CTP), as well as flow-limiting stenoses (*FFR*_*sim*_ *<* 0.8) at some of the untreated sites, specifically the distal LAD, distal RCA and mid LCX. The hypothetical scenario where all these residual flow-limiting lesions are revascularized, indicated in Figure 7 as “optimized”, shows higher restoration in MBF, vastly reduced mass at ischemic risk, as well as the resolution of perfusion defects in almost all the cardiac segments. In the context of a prospective use of the Virtual PCI method, therefore, this specific PCI procedure would be the recommended one based on its predicted hemodynamic outcomes.

## IV. DISCUSSION

This study presents the Virtual PCI method, an operative planning tool based on finite element modelling to predict the hemodynamic outcomes of Percutaneous Coronary Intervention.

While many computational studies focused on structural simulations to optimize stent deployment [28], only a handful investigate the effects on the post-operative hemodynamics, which is extremely important to assess whether the executed intervention is successful in preventing ischemia or it is insufficient. Compared to previous works that focus only on epicardial artery hemodynamics to predict post-operative FFR [5]–[8], the Virtual PCI method provides for the first time an integrated analysis including the 3D map of stress Myocardial Blood Flow in addition to FFR.

This is enabled by our use of a multiphysics model with a 3D description of myocardial perfusion, which provides several advantages over epicardial Computational Fluid Dynamics alone. Firstly, it allows for a consistent association between ischemic regions and specific segments in the epicardial coronary tree, enabling a more informed screening for multiple, possible revascularization options. Secondly, it allows for a direct investigation of regional myocardial ischemia and its evolution after the intervention. Finally, it enables a complete model personalization by exploiting pre-intervention stress CTP images to precisely calibrate its parameters: starting from a calibration method we already proposed in [10], in this work we expand it to provide, for the first time, a pointwise calibration strategy for microcirculation parameters, e.g. the local density of microvessels. This is a crucial improvement for the interpretability of the computational results, allowing cardiologists to interpret simulation outputs in the same way they do with stress CTP imaging.

Our hemodynamic results indicate that the choice of the number and location of stenotic sites to treat has major effects not only in terms of the overall clinical outcome (as already documented [29]) but also in terms of the specific hemodynamic state achieved after PCI, potentially spotting insufficient restoration in MBF (residual ischemia) or the insurgence of additional flow-limiting stenoses at some untreated stenotic sites. Since these effects occur only after some stenotic sites are revascularized, these findings confirm that the outcomes of PCI procedures cannot be predicted based only on pre-treatment imaging and require a functional simulation of the post-intervention scenario instead. Moreover, in our comparative study we show that the Virtual PCI approach holds great potential in treatment screening for the identification of the optimal procedure to restore MBF across the entire myocardium. All these results strongly support the use of the proposed Virtual PCI method for the optimization of PCI procedures.

The results of our validation study show that the Virtual PCI method is effective in predicting the evolution of perfusion defects and in quantifying the post-PCI mass at risk of inducible ischemia, particularly when the pre-intervention percentage of mass at risk is low or moderate. Indeed, we observe a high agreement between the computational results and the stress CTP imaging at follow-up, which is used as validation benchmark. From a clinical perspective based on a 17-segment evaluation, the Virtual PCI method shows a good performance in the detection of regional perfusion defects, with a perfect accordance with follow-up stress CTP in 7 out of 11 evaluated macro-territories (territories attributed to one of the three major coronaries) across all the patients. Performance in the localization of ischemia is also evaluated, showing however a lower accuracy, where perfusion defects may be attributed to adjacent segments with respect to the ones documented as ischemic by stress CTP. Importantly, we do not observe any macro-territory where a perfusion defect documented by stress CTP is completely missed by the Virtual PCI method.

From a clinical perspective, the Virtual PCI approach can be effectively integrated in currently established workflows to manage patients affected by obstructive CAD, providing incremental prognostic and therapeutic value, due to some key features.

Firstly, it is based only on noninvasive scans, namely cCTA at rest and stress CTP. These exams are often performed together in a sequential protocol [17], [30] to integrate the high negative predictive power of cCTA [31] with the additional functional information on MBF provided by stress CTP. Our approach, therefore, can be used at an early stage, screening for PCI treatment options much earlier than when the patient access the operative room. Even though the total time required for the whole computational analysis is on the higher side, consisting of *≃* 3 hours of operational time to generate the anatomical model and additional *≃* 3 hours of simulation time for each simulated treatment option, this is still compatible with the time frame of elective revascularization, which is usually scheduled a a few days after the CT scans. In any case, current research on how to improve efficiency is under investigation.

Secondly, simulation outputs reproduce quantities that are already part of the clinical practice with validated cutoffs, namely the FFR index for the hemodynamic significance of coronary lesions [2], [3] and the stress MBF for the assessment of inducible ischemia [18], enabling a direct interpretation of computational outcomes.

Finally, the use of stress MBF for model calibration, as opposed to FFR, enables the use of our method also in patients that have already been revascularized with stenting. These patients, which cannot be analyzed with existing FFR-based PCI planning tools, represent a relevant group often showing a delicate clinical condition and would greatly benefit from optimized revascularization strategies.

This work presents some limitations:

- we used stress CTP imaging at a follow up of 5-6 years for validation, whereas the Virtual PCI method is designed to predict the hemodynamic outcomes right after the procedure. Although we minimized the impact of this discrepancy by analyzing only patients who exhibited stable atherosclerosis (i.e. without relevant disease progression), a comparison with perfusion imaging acquired shortly after PCI would be beneficial for confidence in validation;
- although the analyzed patients show vastly different clinical conditions and a relevant number (*m* = 15) of lesions revascularized, the population size (*n* = 6) defines this work as a proof-of-concept study and a larger population is required to draw meaningful conclusions regarding the accuracy of the Virtual PCI method. Still, we believe the present work to be an essential step to define the methodological framework and a precise workflow integrated with the clinical practice.

## V. CONCLUSION

The Virtual PCI tool presented in this study constitutes a consistent computational framework to predict the hemodynamic outcomes of PCI procedures and can be effectively integrated into clinical practice at an early stage in the patient management flow, exploiting the established imaging protocol of rest cCTA + stress CTP acquisition. When compared to stress CTP at follow-up in patients treated with elective PCI, our method shows good performances in predicting the evolution of perfusion defects and in the quantification of the cardiac mass at ischemic risk after revascularization. Also, the clinical interpretation of the Virtual PCI predictions is comparable to the interpretation of stress CTP at follow-up, further corroborating its predictive power. Still, we recognize the need for a larger retrospective study to draw meaningful conclusions regarding the method’s accuracy and to clearly define its scope of applicability. The Virtual PCI method shows potential for its prospective use, showing relevant differences in the hemodynamic outcomes of different potential revascularization strategies on the same patient, as well as proving capable of identifying the optimal strategy in a complex clinical case. We believe that the comprehensive hemodynamic analysis provided by the Virtual PCI method can lead to more effective strategies to guide PCI procedures by introducing innovative criteria for treatment selection, for example the minimum intervention that guarantees restoration of MBF across the entire myocardium.

## Data Availability

All data produced in the present study are available upon reasonable request to the authors

## ACKNOWLEDGMENT

GMP has been supported by Bracco Imaging S.p.A., by Consiglio Nazionale delle Ricerche (CNR) and by Italian PNRR research funding, Missione 4, DM226/2021.

GMP and CV are members of the INdAM group GNCS “Gruppo Nazionale per il Calcolo Scientifico” (National Group for Scientific Computing).

CV has been partially supported by: i) the European Union-Next Generation EU, Mission 4, Component 1, CUP: D53D23018770001, under the research project MIUR PRIN22-PNRR n.P20223KSS2, “Machine learning for fluid structure interaction in cardiovascular problems: efficient solutions, model reduction, inverse problems”, ii) the Italian Ministry of Health within the PNC PROGETTO HUB LIFE SCIENCE - DIAGNOSTICA AVANZATA (HLS-DA) “INNOVA”, PNCE3-2022-23683266–CUP: D43C22004930001,

within the “Piano Nazionale Complementare Ecosistema Innovativo della Salute” Codice univoco investimento: PNCE3-2022-23683266; iii) the Italian research project MIUR PRIN22 n.2022L3JC5T “Predicting the outcome of endovascular repair for thoracic aortic aneurysms: analysis of fluid dynamic modelling in different anatomical settings and clinical validation” ; iv) Italian Ministry of Health within the project “CAL.HUB.RIA” - CALABRIA HUB PER RICERCA INNOVATIVA ED AVANZATA. Code: T4-AN-09, CUP: F63C22000530001.

We acknowledge the CINECA award under the ISCRA initiative, for the availability of high performance computing resources and support, in the ISCRA-C project Vr-PCI.

This study was supported by the European Union– Next Generation EU – NRRP M6C2 – Investment 2.1 Enhancement and strengthening of biomedical research in the NHS-Project: PNRR-POC -2022-12376500, CONCERTO.

